# Improved estimation of time-varying reproduction numbers at low case incidence and between epidemic waves

**DOI:** 10.1101/2020.09.14.20194589

**Authors:** Kris V Parag

**Affiliations:** MRC Centre for Global Infectious Disease Analysis, Imperial College London, London, W2 1PG, UK

**Keywords:** Bayesian filters, reproduction numbers, epidemic models, COVID-19, infectious diseases

## Abstract

We construct a recursive Bayesian smoother, termed EpiFilter, for estimating the effective reproduction number, R, from the incidence of an infectious disease in real time and retrospectively. Our approach borrows from Kalman filtering theory, is quick and easy to compute, generalisable, deterministic and unlike many current methods, requires no change-point or window size assumptions. We model R as a flexible, hidden Markov state process and exactly solve forward-backward algorithms, to derive R estimates that incorporate all available incidence information. This unifies and extends two popular methods, EpiEstim, which considers past incidence, and the Wallinga-Teunis method, which looks forward in time. We find that this combination of maximising information and minimising assumptions significantly reduces the bias and variance of R estimates. Moreover, these properties make EpiFilter more statistically robust in periods of low incidence, where existing methods can become destabilised. As a result, EpiFilter offers improved inference of time-varying transmission patterns that are especially advantageous for assessing the risk of upcoming waves of infection in real time and at various spatial scales.

**Author Summary:** Inferring changes in the transmissibility of an infectious disease is crucial for understanding and controlling epidemic spread. The effective reproduction number, R, is widely used to assess transmissibility. R measures the average number of secondary cases caused by a primary case and has provided insight into many diseases including COVID-19. An upsurge in R can forewarn of upcoming infections, while suppression of R can indicate if public health interventions are working. Reliable estimates of temporal changes in R can contribute important evidence to policymaking. Popular R-inference methods, while powerful, can struggle when cases are few because data are noisy. This can limit detection of crucial variations in transmissibility that may occur, for example, when infections are waning or when analysing transmissibility over fine geographic scales. In this paper we improve the general reliability of R-estimates and specifically increase robustness when cases are few. By adapting principles from control engineering, we formulate EpiFilter, a novel method for inferring R in real time and retrospectively. EpiFilter can potentially double the information extracted from epidemic time-series (when compared to popular approaches), significantly filtering the noise within data to minimise both bias and uncertainty of R-estimates and enhance the detection of salient changepoints in transmissibility.

## Introduction

During an unfolding epidemic, one of the most commonly available and useful types of surveillance data is the daily (or weekly) number of newly reported cases. This time-series of case counts, also known as the incidence curve, not only measures the epidemic size and burden, but also provides information about trends or changes in its transmissibility [1], [2]. These trends are captured by the time-varying effective or instantaneous reproduction number, denoted *R*_*s*_ at time *s*, which defines how the number of secondary cases generated per primary case varies across the outbreak [3]. Broadly, when *R*_*s*_ > 1 we can expect and prepare for growing incidence, whereas sustained *R*_*s*_ < 1 signifies that the epidemic is waning and likely to enter a more controlled phase [4].

Inferring changes in *R*_*s*_ given an observed incidence curve is therefore crucial, both to understanding transmissibility and to forecasting upcoming case loads, especially for an ongoing epidemic, where it can help inform policymaking and intervention choices or predict healthcare demands [1], [5]. Real-time and retrospective *R*_*s*_ estimates have been used to characterise rates and patterns of spread in various diseases such as malaria [6] and Ebola virus disease [7]. Such estimates have proven valuable throughout the COVID-19 pandemic, providing updating synopses of global transmission [8] and evidencing the impact of past control actions (e.g. lockdowns and social distancing) [9] or the likelihood of a resurgence in infections when those controls are relaxed [10].

Most studies that infer *R*_*s*_ or related quantities either apply the Wallinga-Teunis (WT) method [2] or the Cori *et al* method, known as EpiEstim [11]. Both methods take complementary viewpoints on how incidence data inform on transmissibility and hence have diverse use-cases. The WT method reconstructs the average number of new cases caused by infectious individuals at *s* and so requires incidence data beyond time *s* for its estimate. It computes the case or cohort reproduction number, 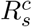, which is a function of *R*_*s*+*j*_ for future times *j* ≥ 0, and is suited for retrospective analyses [12]. Alternatively, EpiEstim infers how past infections propagate to form the incidence observed at *s*, only requiring data prior to time *s*. EpiEstim directly computes instantaneous reproduction numbers, *R*_*s*_, and is preferred for real-time investigations [3].

While both methods provide useful and important estimators of transmission, they are not perfect. Two main limitations exist. First, each suffers from data censoring or edge-effects [3]. Because the WT method is forward-looking i.e., depends on data later than *s*, its estimates are right censored when *s* is close to the last observed time point [12]. In contrast, EpiEstim looks backward in time and suffers edge-effects when *s* is near the first observed time point [11]. Estimates in the vicinity of the start and end of the incidence time-series are therefore unreliable under EpiEstim and the WT method, respectively. Techniques have been proposed to limit this unreliability [5], [13], but the problem is intrinsic, and inevitable near the actual start and end times of an epidemic, where there is necessarily no or sparse data.

This leads into the second key limitation: inference in periods of small incidence. This presents a fundamental challenge for any *R*_*s*_ estimation approach and effectively creates additional edge-effects. When few or no case counts are available to constrain inference, methods are largely driven by their inherent prior distributions and assumptions. This can result in misleading or unreliable estimates and mask subcritical (i.e., *R*_*s*_ < 1) transmission patterns [11], [15]. Small incidence may occur during sustained periods of interventions, when pathogens invade new susceptible regions and also naturally arise by division when analyses are performed at smaller spatial scales (e.g., regional or community levels). Understanding how to best mediate the trade-off between prior assumptions and data when incidence is small is of both statistical and epidemiological significance.

Following a period of low incidence, two important outcomes are possible: either the epidemic continues to exhibit small or zero case counts until it goes extinct, or a resurgence in infections, also termed a second wave, occurs. Inferring, in real time, which of these conditions is likely presents a key challenge for infectious disease epidemiology given the information bottleneck at low incidence [16]. Better inference of transmission under these conditions is currently considered central to designing data-informed COVID-19 intervention exit or relaxation strategies [17]. With many countries facing multiple resurgent waves in this pandemic, estimating fluctuations in transmission during suspected epidemic troughs could be essential to achieving sustained control [10].

Here we present and develop a novel method, termed EpiFilter, for reliably estimating *R*_*s*_ in real time, which ameliorates the above limitations. We take an engineering inspired approach and construct an exact, recursive and deterministic (i.e., EpiFilter produces the same output for fixed input data and requires no Monte Carlo steps) inference algorithm that is quick and easy to compute both across an unfolding outbreak and in retrospect. Our method solves what is called the smoothing problem in control engineering [14]. This means we compute instantaneous reproduction number, *R*_*s*_, estimates that formally integrate both forward and backward looking information. This unifies the WT method and EpiEstim, and largely nullifies their edge-effect issues.

Further, EpiFilter only makes a minimal Markov assumption for *R*_*s*_, which allows it to avoid the strong prior window size and change-point assumptions that existing methods may apply to infer shifts in transmission [9], [11]. Using simulated and empirical data, we show that EpiFilter accurately tracks changes in *R*_*s*_ and provides reliable one-step-ahead incidence predictions. Moreover, we find that EpiFilter is appreciably more robust and statistically efficient than even optimised versions of EpiEstim [13]. Specifically, it does not easily destabilise when performing real-time inference in periods of low incidence, such as in lulls between epidemic waves, and generally it minimises the mean squared error of *R*_*s*_ estimates, while maintaining good coverage and predictive performance.

We illustrate the practical utility of EpiFilter on the COVID-19 incidence curve of New Zealand, which exhibits a second wave that was seeded during a prolonged low incidence period. We find stark improvements in the transmission patterns EpiFilter uncovers. Our method, which is outlined in Fig. 1, provides a straightforward yet formally optimal (in mean squared error sense [18]) solution to real-time and retrospective instantaneous reproduction number estimation. Because it couples minimal prior assumptions with maximum information extraction, it more gracefully handles periods with scarce data. Hopefully our approach will serve as a useful inference tool for investigating the risk of resurgence in COVID-19 and other epidemics. Matlab and R implementations of EpiFilter are available at https://github.com/kpzoo/EpiFilter and its mechanics are explored and validated in the S1 Appendix.

**Fig. 1:**
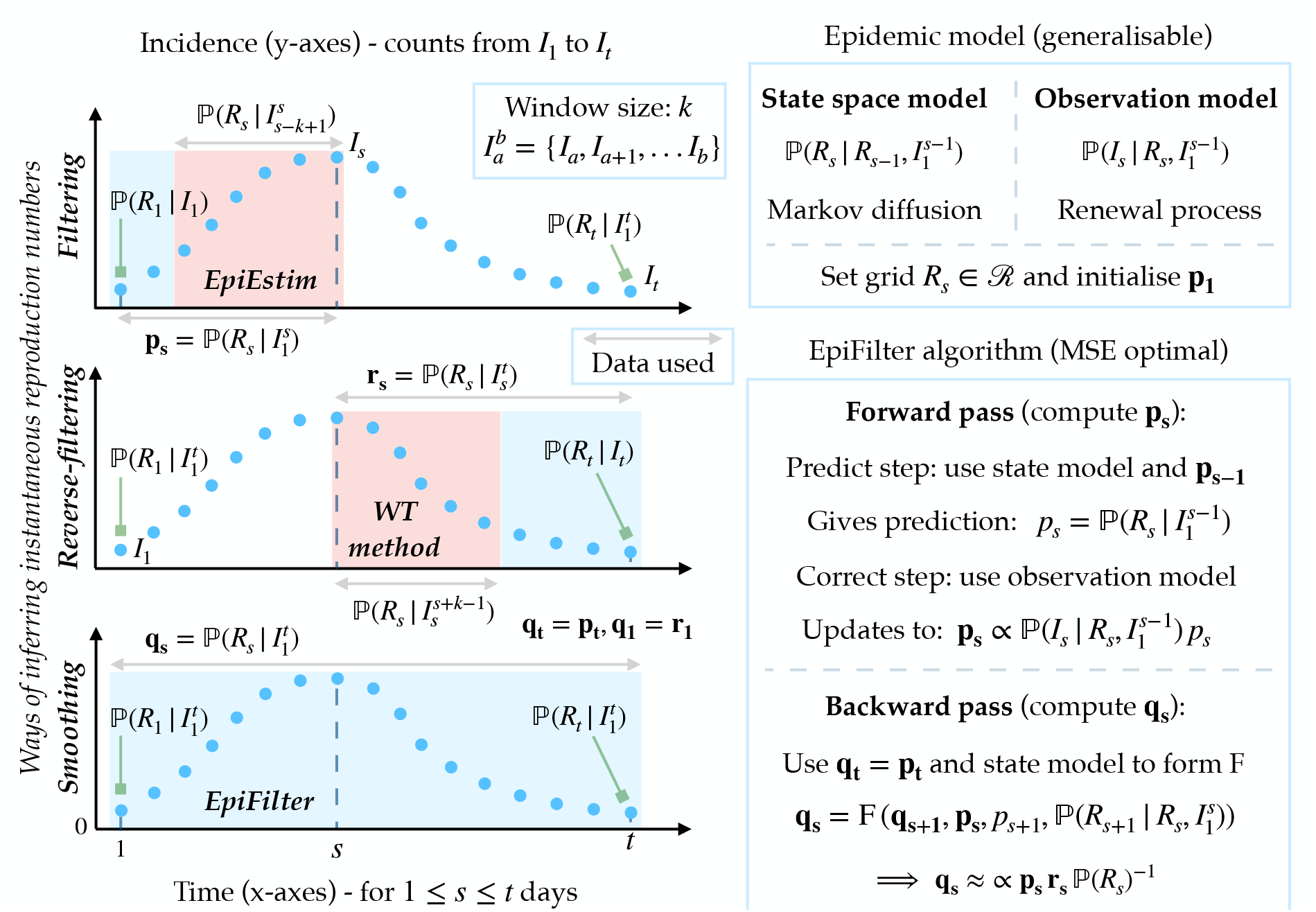
EpiFilter algorithm and relationship to other methods. In the left panels we consider three ways of inferring the instantaneous or effective reproduction number at time *s, R*_*s*_, from the incidence curve, 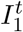 (blue dots). The filtering solution produces the posterior distribution **p**_**s**_ from all data prior to time *s*. EpiEstim approximates this solution by using the subset of data in a window of size *k* into the past. Reverse-filtering considers the complementary part of the incidence curve, leading to **r**_**s**_, which utilises data beyond *s*. The WT method, with future window *k*, approximates this type of solution. Smoothing uses all information from 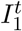to generate **q**_**s**_, which is precisely computed by EpiFilter. Blue windows show the portions of 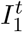 that inform on *R*_*s*_ for each of **p**_**s**_, **r**_**s**_ and **q**_**s**_ while red windows highlight the subsets used by EpiEstim and the WT method. Double arrows indicate data used for constructing various posterior distributions, while square arrows pinpoint instances of those distributions at the edges of 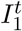. In the right panels we summarise the construction of EpiFilter. We outline the main assumptions (the model box) and computations (the algorithm box) necessary for realising EpiFilter, which allow us to obtain the most informative (and minimum MSE) smoothing posterior distribution **q**_**s**_. See the main text for the specific equations employed in our implementation [14].

## Methods

### Renewal models and inference problems

We consider an infectious disease epidemic observed over some time period 1 ≤ *s* ≤ *t* in a homogeneous and well-mixed population. While epidemics actually spread on dynamic networks involving stratified contact structures, homogeneous models can provide useful real-time insight into key transmission patterns and are more easily fit and verified with routine surveillance data such as incidence curves [19]. If the incidence or number of newly infected cases at time *s* is *I*_*s*_ then the set 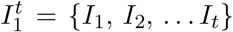 is the incidence curve of the epidemic. We assume that incidence is available on a daily scale so that 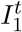 is a vector of *t* daily counts but weeks or months could be used instead. A common problem in infectious disease is the inference of the transmissibility of the epidemic given this curve. The renewal model [1], [20] presents a general and popular framework for investigating this problem and its estimates, in some instances, can even approximate those from detailed network models [21].

The renewal model posits that epidemic transmissibility, summarised by the effective or instantaneous reproduction number, *R*_*s*_, generates the observed incidence as in Eq. (1). We assume incidence counts are Poisson distributed with Pois indicating Poisson noise. Here 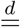 signifies equality in distributions and | means ‘conditioned on’.

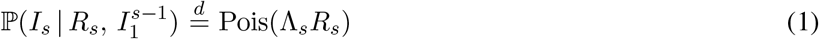

While Eq. (1) does not directly model how susceptible individuals become infected, these effects are encoded in the reproduction number *R*_*s*_, which measures the secondary cases generated per effective primary case at *s* [3]. The quantity 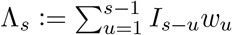, known as the total infectiousness, counts how many effective past cases are still infectious at *s* i.e., it describes the number of circulating cases that can actively transmit.

The generation time distribution of the epidemic, 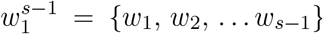, controls how past incidence influences Λ_*s*_, with *w*_*u*_ as the probability that a primary case takes between *u*−1 and *u* days to generate a secondary case [1]. We make the standard assumption that 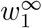 is well approximated by the serial interval distribution of the epidemic of interest, which is known [11]. The serial interval is the time between symptom onset of a primary and its secondary case. While onset times are more practically measurable than actual infection times they do not include asymptomatic or subclinical cases. This approximation can limit inference (for example, serial intervals often have larger variances than generation times), but methods are being developed to improve its quality [22].

We focus on inferring the complete set of *R*_*s*_ values, denoted 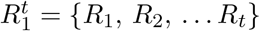, given the incidence curve 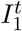 with *t* as the last recorded time. Estimating 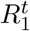 is important because changes in the values of instantaneous reproduction numbers often signify key transitions in epidemic transmissibility, which might be due to the imposition or relaxation of interventions. Instantaneous reproduction numbers are also the basis of other transmissibility metrics, such as cohort reproduction numbers or growth rates [3]. We define three main inference problems, based on how information in 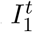 is recruited to construct every *R*_*s*_ estimate. We represent these problems in terms of the posterior distribution their solutions induce over possible *R*_*s*_ values. Estimates are functions of these posteriors.

The first is called filtering, where we sequentially compute the filtering posterior 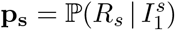 for every *s* ≤ *t* [23]. Filtering only uses incidence data up to time *s*, 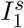, for inferring *R*_*s*_. Solving this problem is fundamental to real-time inference [24]. Filtering solutions are commonly employed for inferring instantaneous reproduction numbers. The second problem, which we call reverse-filtering, is the complement of the first. The reverse-filtering posterior 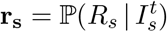, is important for retrospective or backward-looking estimates and infers *R*_*s*_ from incidence beyond *s* i.e., 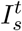 [25]. *Practical R*_*s*_ calculations do not involve reverse-filtering. Instead, the information used by **r**_**s**_ is implicit to deriving cohort reproduction numbers, 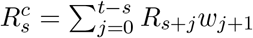 [12]. Later we show that estimates from EpiEstim and the WT method are related to the **p**_**s**_ and **r**_**s**_ distributions, respectively.

The last problem, which is termed smoothing, is our main interest. It asks the question: how can we construct an *R*_*s*_, at every *s* ≤ *t*, that integrates all available incidence information from 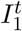. To solve this problem we must formulate the smoothing posterior distribution 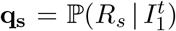 [14]. Functions of this posterior would then yield maximally informed instantaneous reproduction number estimates (and cohort reproduction number estimates by extension). Note that **p**_**s**_, **r**_**s**_ and **q**_**s**_ depend on the choices of state space model, which describes the dynamics of *R*_*s*_ across time, and observation model, which explains how changes in *R*_*s*_ lead to trends in observed incidence data [18]. These models encode our assumptions about the epidemic of interest and determine how estimates trade off assumptions against data. We next explore how posterior distribution selection determines performance, especially when data are scarce, and examine how EpiEstim and the WT methods fit within this framework.

### Inference methods and low incidence

We mostly detail EpiEstim as instantaneous reproduction number, *R*_*s*_, estimates are the main focus of this work. EpiEstim assumes that the estimate of *R*_*s*_ at time *s* depends on a rolling past window of data, *τ* (*s*) = {*s, s* − 1, …, *s* − *k* + 1}, of size *k* [11]. Consequently, at time *s*, only size *k* subsets of the incidence, 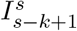, and total infectiousness, 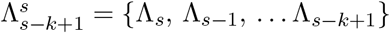, are considered informative about *R*_*s*_. Their sums over this window are *i*_*τ*(*s*)_ = Σ _*u*∈*τ*(*s*)_ *I*_*u*_ and *λ*_*τ*(*s*)_ = Σ _*u*∈*τ*(*s*)_ Λ_*u*_. When *k* = 1, EpiEstim only uses the most recent case count, *I*_*s*_, (and Λ_*s*_) to estimate *R*_*s*_. While this maximises flexibility, it usually results in over-fitting and so larger windows are often employed to trade off estimate variance with bias [11], [26].

EpiEstim therefore effectively solves a filtering problem, as discussed in the previous section. The filtering posterior distribution produced by EpiEstim is restricted to the informative window *τ* (*s*) and denoted 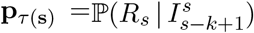. This is parametrised by the shape-scale gamma posterior distribution in Eq. (2).

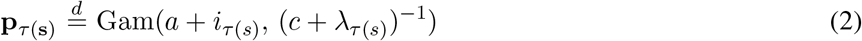

The posterior **p**_*τ* (**s**)_ results from combining a gamma prior distribution on *R*_*s*_, 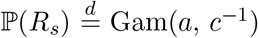 with a Poisson observation likelihood for the incidence (see Eq. (1)), as detailed in [11], [13]. We never explicitly include Λ_*s*_ terms in our notation (e.g., we could write 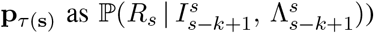 since they appear naturally when using Eq. (1) and the key difference among our inference problems relate to *I*_*s*_ terms.

The posterior mean estimate from Eq. (2) is constructed as 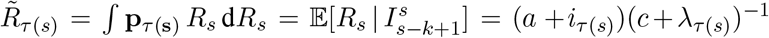. The variance around this estimate is (*a* + *i*_*τ*(*s*)_)(*c* + *λ*_*τ*(*s*)_)^−2^. The observation model is given by Eq. (1) but the state space model of EpiEstim is not explicit. However, if *R*_*τ* (*s*)_ is the assumed average reproduction number in *τ* (*s*) (which is used to estimate *R*_*s*_) then *λ*_*τ*(*s*)_ *R*_*τ*(*s*)_ = Σ _*u*∈*τ*(*s*)_ Λ_*u*_*R*_*u*_ [13]. Thus, EpiEstim somewhat incorporates a linear moving average state space model, and assumes that the filtering distribution **p**_***s***_ *≈* **p**_*τ* (**s**)_ by deeming data outside 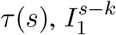, as effectively uninformative [12]. Since **p**_*τ*(**s**)_ can be computed sequentially across an ongoing epidemic, EpiEstim provides real-time inference.

The WT method takes a complementary approach to EpiEstim, computing transmissibility over a forward-looking window *γ*(*s*) = {*s, s* + 1, …, *s* + *k* − 1} [2]. Often *k* = *t* − *s* + 1 i.e., the window extends to the last observed incidence. The WT method uses the observation model of Eq. (1) and has an implicit moving average state model 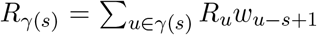, which leads to its cohort reproduction number estimates [3]. As this method effectively uses future information [12], it implicitly involves approximating the reverse-filtering distribution (see previous section) as 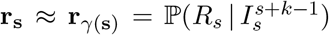. We illustrate the information windows employed by the WT method and EpiEstim, as well as the complete filtering and reverse-filtering windows in Fig. 1. The goodness of the windowed distributions **p**_*τ*(**s**)_ and **r**_*γ*(**s**)_ as approximations to the general posterior distributions **p**_**s**_ and **r**_**s**_ will depend on *k* and the appropriateness of the state model underlying each method [13].

While EpiEstim and the WT methods are powerful tools for inferring transmissibility in real-time and in retrospect, they have two main and related limitations, which necessarily reduce the reliability of their outputs [12]. First, their performance degrades as *s* gets close to 1 for EpiEstim and *t* for the WT method [11]. These edge or censoring effects correspond, at the extreme, to **p**_**1**_ = **p**_*τ* (**1**)_ = ℙ(*R*_1_ | *I*_1_) and **r**_**t**_ = **r**_*γ*(**t**)_ = ℙ(*R*_*t*_ | *I*_*t*_), which are weakly informed posterior distributions. As a result, at the beginning of the incidence curve EpiEstim can be unreliable (and even unidentifiable). The WT method suffers similarly at the end of that curve [1] (see Fig. 1).

The second limitation occurs in phases of the epidemic where incidence is low for a prolonged period of time [17], [26]. In these periods data are sparse and the quality of estimates depend on how well the method of choice mediates between the little available information and its inherent assumptions [15]. We illustrate this with EpiEstim, using the mean and variance of the posterior estimate from Eq. (2), 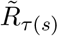, which are defined above. If incidence is small over the window *τ* (*s*) then the sum of incidence, *i*_*τ* (*s*)_, and the total infectiousness *λ* _*τ* (*s*)_ shrink, meaning that the prior hyperparameters, *a* and *c*, strongly influence the resulting estimate mean and variance. This contrasts the data-rich scenario when an epidemic is large, where *i*_*τ* (*s*)_ and *λ* _*τ* (*s*)_ overpower *a* and *c*.

Further, should a sequence of *n* ≥ *k* zero-incidence days occur, then *i*_*τ* (*s*)_ = 0 and *λ* _*τ* (*s*)_ → 0 as *n* increases, with a rate controlled by the serial interval of the epidemic. Here the shape parameter of **p**_*τ* (**s**)_ is exactly that of the prior distribution ℙ(*R*_*s*_). As *λ* _*τ* (*s*)_ decays, **p**_*τ* (**s**)_ → ℙ(*R*_*s*_) (see Eq. (2)), *R*_*s*_ becomes statistically unidentifiable from the window of data and inference is completely prior driven [26], [27]. While lack of data is a fundamental limitation, the point at which we lose inferential power is not fixed, and depends on the window size, *k*. Studies that formally optimised *k* for estimate reliability, found that small *k* is needed to infer sharp changes in transmissibility (e.g. due to lockdowns) [13], indicating that these issues can be acute. Analogous effects occur in the WT method if there are few incident cases across its forward-looking window *γ*(*s*) [12].

These prior-driven scenarios are realistic for epidemics in waning or tail phases, and can precede either elimination (i.e., epidemic extinction) or resurgence [28]. While some estimate degradation is guaranteed for any *R*_*s*_ inference method when faced with either edge-effects or low incidence, robustness can still be improved. Edge-effects can be largely overcome by constructing the smoothed posterior distribution for estimating the instantaneous reproduction number *R*_*s*_, denoted **q**_**s**_. Solving the smoothing problem melds the advantages of the opposite looking windows of EpiEstim and the WT method, removing the vulnerability near the ends of the incidence curve 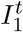. This follows as **q**_**1**_ = **r**_**1**_ and **q**_**t**_ = **p**_**t**_ (see Fig. 1). Further, by maximising the information used for inferring every *R*_*s*_ and by minimising our state model assumptions, we can ameliorate the impact of low incidence. We next develop a method, termed EpiFilter, to realise these improvements.

### Bayesian (forward) recursive filtering

We reformulate the inference problem of estimating instantaneous reproduction numbers *R*_*s*_ from past incidence 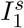 as an optimal Markov state filtering problem. Filtering describes a general class of engineering problems aimed at optimally, usually in a mean squared error (MSE) sense, inferring some hidden state in real time from noisy observations [14], [18]. Given some functions *f*_*s*_ and *g*_*s*_, which describe the state (*R*_*s*_ in our case) space dynamics and the process of generating noisy observations (the *I*_*s*_ here), the filtering problem tries to construct the posterior distribution **p**_**s**_ (see previous section) [23], which EpiEstim approximates. The conditional mean estimate 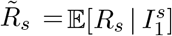 leads to the minimum MSE of 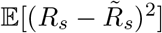 [23], which depends on all the past information.

The famed Kalman filter [24] was the genesis of these methods. Here we focus on Bayesian recursive filters for models with noisy count observations. These generalise the Kalman filter [23] and have been successfully applied to similar problems in phylodynamics and computational biology [29], [30], [31]. We reconsider our renewal model inference problem within this engineering state-observation framework, as described in Eq. (3) [14].

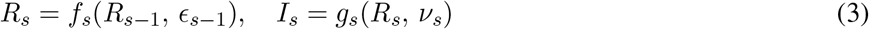

Here *R*_*s*_ is the hidden Markov state that we wish to infer. It dynamically depends on the previous state *R*_*s*−1_ and a noise term *ϵ*_*s*−1_ via *f*_*s*_. Observation *I*_*s*_ is then elicited due to *R*_*s*_ and a noise term *ν*_*s*_, according to *g*_*s*_ [32].

We develop our filter under two very mild assumptions. First, we define some closed space, ℛ, over which *R*_*s*_ is valid. For a given resolution *m*, extrema *R*_min_ and *R*_max_, and grid size *δ*= *m*^-1^(*R*_max_ − *R*_min_) then ℛ := {*R*_min_, *R*_min_ + *δ*, …, *R*_max_}. This means the instantaneous reproduction number *R*_*s*_ must take a discrete value in ℛ, the *i*^th^ element of which is denoted ℛ[*i*]. We formalise this notion in Eq. (4a).

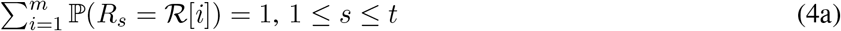

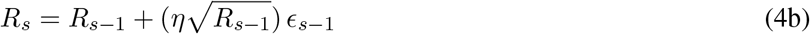

This is not restrictive since we can compute our filter for large *m* if needed and usually we are only interested in *R*_*s*_ on a coarse scale (e.g., policymakers may only want to know if *R*_*s*_ ≤ 1 or not). Other approaches, which depend on MCMC or related sampling methods (e.g., [9] and some implementations of EpiEstim), all implicitly assume some discretisation [29]. In the S1 Appendix (Fig. A1) we show that often convergence occurs at small *m*. Second, we propose a linear model for *f*_*s*_, as defined in Eq. (4b). There *ϵ*_*s*−1_ is a standard white noise term i.e., 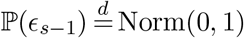 with Norm signifying a normal distribution and *η* as a free parameter. We assume that a noisy linear projection of states over consecutive time-points provides a good approximation of the state trajectory. Not only is this assumption standard in engineering [23] and epidemiology [33] but it is also more flexible than the state model inherent to EpiEstim and the WT method. We scale the noise of this projection by a fraction, *η* < 1, of the magnitude of *R*_*s*−1_. This parameter controls the correlation among successive instantaneous reproduction numbers (and hence the state noise) but ensures *R*_*s*_ is a-priori non-negative.

Our observation model, *g*_*s*_, is implicit and leads to the probability law in Eq. (1). As a result, both our observations and state models are discrete (see Fig. 1 for summaries). Because the state model governing *R*_*s*_ in Eq. (4b) is stochastic, Eq. (1) actually describes an over-dispersed (doubly stochastic) Poisson incidence curve. Consequently, *η*, allows us to better model some of the heterogeneity in transmissibility, and may increase robustness to violations of the well-mixed assumption inherent to renewal models [21]. We can optimise our choice of *η* value by minimising the incidence one-step-ahead predictions that result from our observation model [13].

We now define the Bayesian recursive filtering procedure, which is a main contribution of this work, and can be solved exactly, in real-time and with minimal computational effort. We adapt general recursive filtering equations from [14], [18], [32], [25], which are valid for various types of observation and state models, to our renewal model inference problem. The proof of the equations we employ can be found in these works. While we solve discrete, univariate problems (our state model is one dimensional), extensions to continuous-time, multivariate problems also exist [23], [30]. These recursive equations can also be approximately solved using particle filters [14], [32].

Recursive filtering involves two steps: prediction and correction. The first, given in Eq. (5a), constructs a sequential prior predictive distribution, 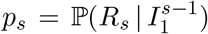. This is informed by past incidence data 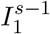 and the last state *R*_*s*−1_. The second step then corrects or updates this prior prediction into a posterior filtering distribution, **p**_**s**_, which constrains *p*_*s*_ using the latest observation, *I*_*s*_, according to Eq. (5b).

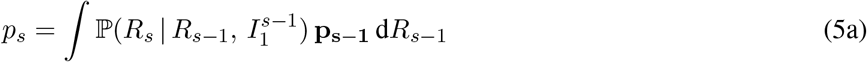

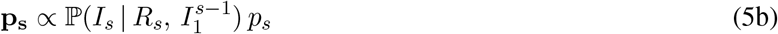

Here 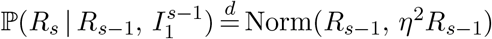 is the state model from Eq. (4b), 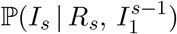 is the observation model from Eq. (1) and the constant of proportionality for Eq. (5b) is simply a normalising factor.

Solving Eq. (5) iteratively and simultaneously over the grid of ℛ leads to our novel real-time estimate of the time-varying effective reproduction number. We initialise this process with a uniform prior distribution over ℛ for **p**_**1**_ and note that *p*_*s*_ and **p**_**s**_ are *m* element vectors that sum to 1, with *i*^th^ term corresponding to when *R*_*s*_ = ℛ[*i*]. Eq. (5) forms the first half of EpiFilter, is flexible and can be adapted to many related problems [14]. A key difference between EpiFilter and the EpiEstim-type methods [11], [13] is that the latter approximate the distributions 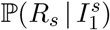 and 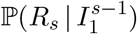 with 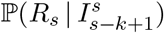 and ℙ(*R*_*s*_), respectively. Estimators based on these approximations can be suboptimal, especially when data (i.e., cases) are scarce.

### Bayesian (backward) recursive smoothing

While Eq. (5) provides a complete real-time solution to the filtering problem, it is necessarily limited at the starting edge of the incidence curve, where past data are sparse or unavailable. Further, because it does not update past estimates as new data accumulate, it cannot provide optimal retrospective estimates. Here we develop the second half of EpiFilter, which involves solving the optimal smoothing problem i.e., computing the smoothing posterior distribution 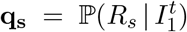, which provides maximally informed estimates of the instantaneous reproduction number *R*_*s*_, given the complete incidence curve 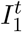. To our knowledge, smoothing has not yet been explicitly considered in infectious disease epidemiology (either exactly or approximately).

We specialise the general methodology from [14], [25] to obtain the recursive smoother of Eq. (6a). This equation uses the filtering distribution, 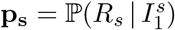 and the predictive distributions 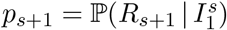, which we obtain from Eq. (5). Our state model means that 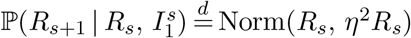.

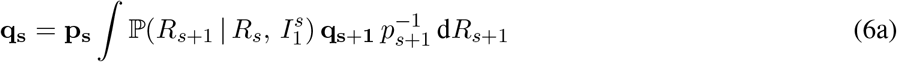

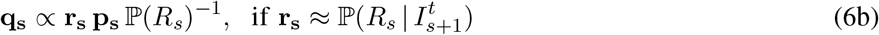

We realise Eq. (6a) exactly by taking a forward-backward algorithmic approach (this is the backward pass whereas Eq. (5) is the forward one). We solve this equation by noting that **q**_**t**_ = **p**_**t**_ and iterating backwards in time to obtain the first smoothing distribution **q**_**1**_. The integrals become sums over our grid ℛ and distributions are *m* element vectors. Eq. (6a) sequentially updates our earlier filtering solutions to include future data, forms the second half of EpiFilter and can also be approximately solved using particle smoothers [14].

This approach neatly links the filtering and smoothing distributions **p**_**s**_ and **q**_**s**_. If we assume that the reverse-filtering distribution **r**_**s**_ is reasonably approximated by 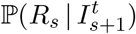 then we can also apply the two-filter smoothing solution of [25] to get Eq. (6b). If either future 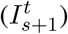 or past 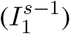 incidence is uninformative then either **r**_**s**_ or **p**_**s**_ will reduce to the prior ℙ(*R*_*s*_), leading to **q**_**s**_ ∝ **p**_**s**_ or **q**_**s**_ ∝ **r**_**s**_, respectively. The end and beginning of the epidemic provide important examples of each of these scenarios. Consequently, Eq. (6b) shows how smoothing connects EpiEstim and the WT methods, and explains why EpiFilter, which can be used for both real-time and retrospective inference, better overcomes edge-effects and periods of low data.

Further, the smoothed posterior **q**_**s**_ yields the conditional mean estimate 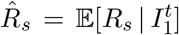, which is known to significantly improve on the MSE of the filtered equivalent 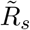 (see previous section) [23]. While filtering provides the minimum MSE estimator of every instantaneous reproduction number *R*_*s*_ given past knowledge, smoothing provides the minimum given all knowledge. This relationship is formal, with filtered and smoothed MSE values mapping to the amount of mutual information that 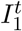 provides about 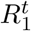 [34]. Extracting the maximum information from the incidence curve 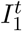 should engender estimates that are more robust and statistically efficient in periods of low incidence. We summarise the EpiFilter algorithm in Fig. 1.

While our main interest is on optimised and rigorous real-time and retrospective estimates of transmissibility, which are completely defined by the smoothing distribution **q**_**s**_, we may also want to predict future incidence, for informing epidemic preparedness plans and for validating past *R*_*s*_ estimates [13], [35]. We compute the filtered one-step-ahead posterior predictive distribution as in Eq. (7) (integrals are over ℛ) [14].

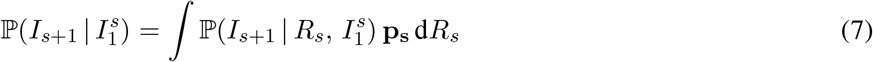

We assume, as in [36], that 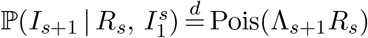. Replacing **p**_**s**_ with **q**_**s**_ yields the smoothed equivalent of Eq. (7). We will use Eq. (7) to compare EpiFilter against APEestim, which is the prediction-optimised version of EpiEstim, developed in [13]. Since predictions depend strongly on **p**_**s**_ or **q**_**s**_, optimising these distributions can be important, for example, when forecasting second waves of infection.

Last, we comment on how we represent uncertainty in our estimates and predictions. Often we will provide 95% equal tailed Bayesian credible intervals. These are computed directly from the 2.5^th^ and 97.5^th^ quantiles of the relevant posterior distribution and used to assess performance statistics such as coverage of true values.

## Results

### Improved estimation at low incidence

The reliable estimation of time-varying effective reproduction numbers, *R*_*s*_, at low incidence, *I*_*s*_, is a key challenge limiting our understanding of transmission [17]. Periods with small counts of new cases contain little information and so present necessary statistical difficulties [26]. Here we compare EpiFilter, which allows exact inference over a state grid ℛ, with EpiEstim and APEestim at these data-poor settings. We use EpiEstim with weekly and monthly windows. Weekly windows are the default recommendation in [11]. We include monthly ones as long windows can improve robustness at low incidence [28]. APEestim, developed in [13], optimises the window choice of EpiEstim to minimise one-step-ahead prediction errors. The assumptions and choices inherent in estimation methods become important and visible when data are scarce, and can bias inference or support spurious predictions [15].

We generate epidemics using the Poisson noise model of Eq. (1) under the serial interval distribution of Ebola virus disease described in [37]. We examine three diverse scenarios in Fig. 2. These describe (A) rapidly controlled epidemics (*R*_*s*_ step changes from 2 to 0.5 at *s* = 100), (B) small outbreaks with exponentially rising and falling *R*_*s*_ (change-point at *s* = 30 and rates of 0.02 and -0.008 per time unit) and (C) medium outbreaks that are initially controlled (*R*_*s*_ changes from 4 to 0.6 at *s* = 40) then resurge (*R*_*s*_ rebounds to 2 at *s* = 80) into large epidemics, before finally being suppressed (*R*_*s*_ = 0.2 from *s* = 150). These scenarios are similar to ones investigated in [12], [28] and describe various epidemic dynamics that culminate in elimination. We simulate 200 incidence curves for each scenario and apply APEestim (optimal window *k*^∗^), EpiEstim (*k* = 7 and 31) and EpiFilter (state noise *η* = 0.1) to estimate *R*_*s*_ and sequentially predict *I*_*s*_ (given data up to *s* − 1) for each curve.

**Fig. 2:**
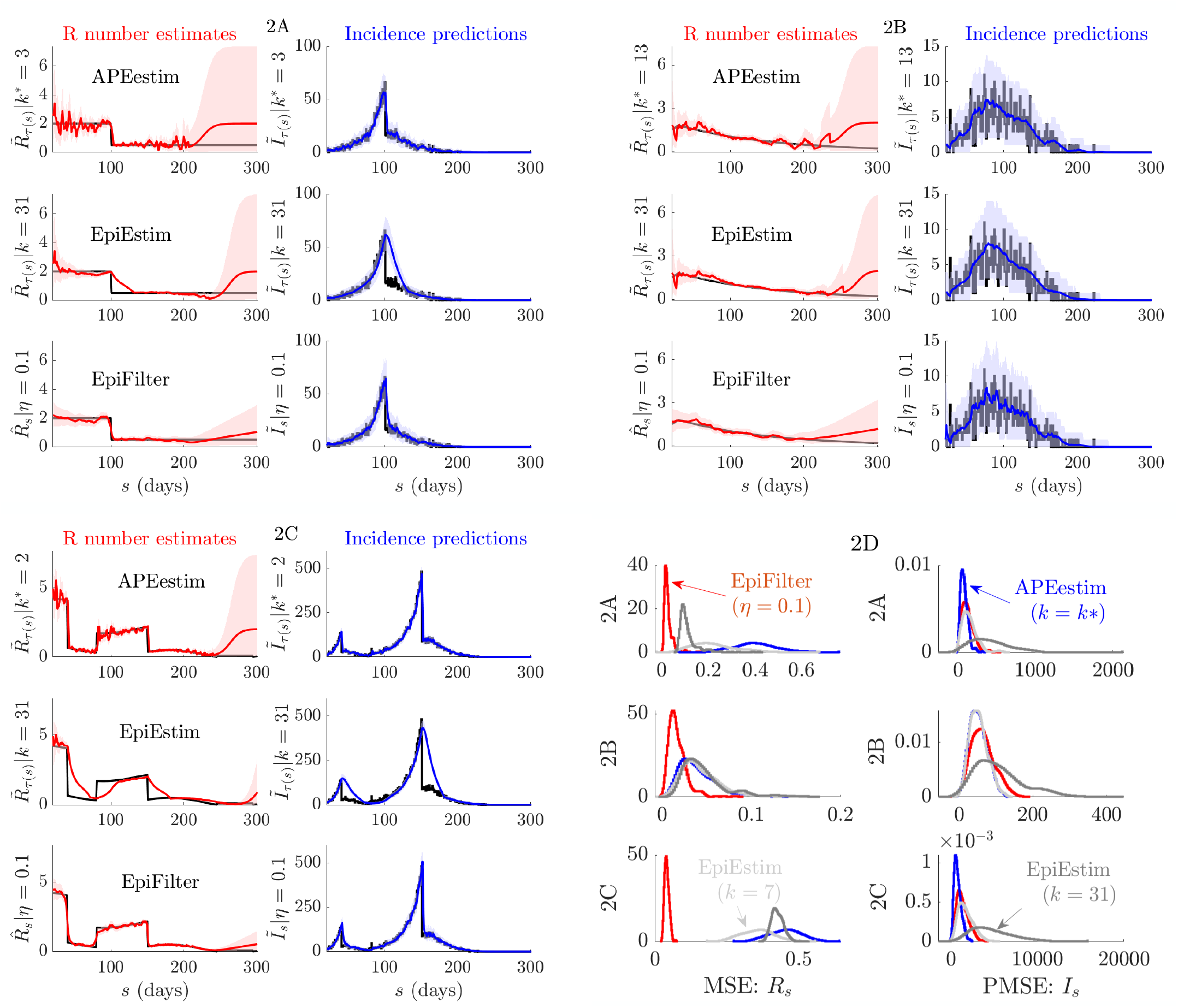
Small or waning epidemics. We compare reproduction number estimates (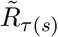 or 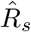) and one-step-ahead incidence predictions (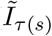 or 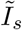) from APEestim with optimal window *k*^∗^, EpiEstim with window *k* and EpiFilter with state noise *η*. We simulate 200 epidemics with low daily case numbers or long tails (long sequences of zero cases) using the standard renewal model (Eq. (1)) for three scenarios, representative examples of which are given in A-C. The true *R*_*s*_ and *I*_*s*_ are in black. All mean estimates or predictions are in red and blue with 95% credible intervals. APEestim and EpiEstim use a Gam(1, 2) prior distribution and EpiFilter a grid with *m* = 2000, *R*_min_ = 0.01 and *R*_max_ = 10. In D we provide statistics of the MSE of these estimates (relative to *R*_*s*_) and the PMSE of these predictions (relative to *I*_*s*_) for all 200 runs. We find that EpiFilter is more robust to small incidence (better uncertainty), whereas the other approaches can quickly decay to their prior distribution. It achieves significantly smaller MSE (2-10 fold reductions) and comparable PMSE to APEestim (which is optimised for prediction).

For the first two methods we compute mean one-step-ahead incidence predictions 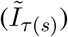 as in [13] and instantaneous reproduction number estimates 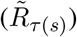 from Eq. (2) with *τ* (*s*) delimiting the window times used. We obtain smoothed EpiFilter estimates 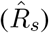 and filtered predictions 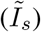 from Eq. (6) and Eq. (7). We could use smoothed predictions 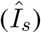, which implicitly include data beyond *s* − 1, to assess fit but as we want to test model adequacy 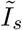 is more appropriate [38]. Our main focus is on real-time performance so we do not investigate the WT method. See [11], [12] for comparisons of the WT method and EpiEstim. Panels A-C of Fig. 2 provide representative single runs (we only show *k* = 31 for EpiEstim as *k* = 7 is often similar to APEestim) while panel D presents reproduction number MSE and one-step-ahead predicted incidence MSE (PMSE) distributions from the 200 runs.

In the Methods we showed that since EpiEstim-type methods group data over some window into the past, they revert to their prior distribution as the total cases in this window becomes small. During low incidence periods, e.g., when the epidemic is waning, using long windows makes sense [28]. However, this reduces predictive accuracy as fluctuations in *R*_*s*_ are underfit. This is the use-case for APEestim, which optimises for prediction. The consequences of these trade-offs are made clear in A-C of Fig. 2 where APEestim provides the best one-step-ahead *I*_*s*_ predictions but quickly reverts to its prior (seen as wide credible intervals) when cases are few. Longer window EpiEstim improves on this destabilisation but cannot track salient fluctuations in *R*_*s*_. Panel D of Fig. 2 verifies these trends. APEstim has the smallest PMSE but often worse MSE than the larger *k* EpiEstim choices.

Interestingly, EpiFilter is able to jointly optimise both instantaneous reproduction number tracking and incidence predictions (computed via Eq. (7)). In A-C of Fig. 2 we see that EpiFilter maintains stable and accurate estimates of *R*_*s*_ and only slowly reverts to its prior (which has the same support as that of EpiEstim and APEesim). However, unlike long window methods it does not sacrifice prediction fidelity. The improvement in MSE shown in panel D of Fig. 2 is stark (the numerical reduction in MSE when compared to the next best method is on average at least 2-fold and often 10-fold). The PMSE, while larger than that of APEestim (which is optimised for predictions) is still good. These points are reinforced in Fig. 3, which expands on the statistics of scenarios A-C.

**Fig. 3:**
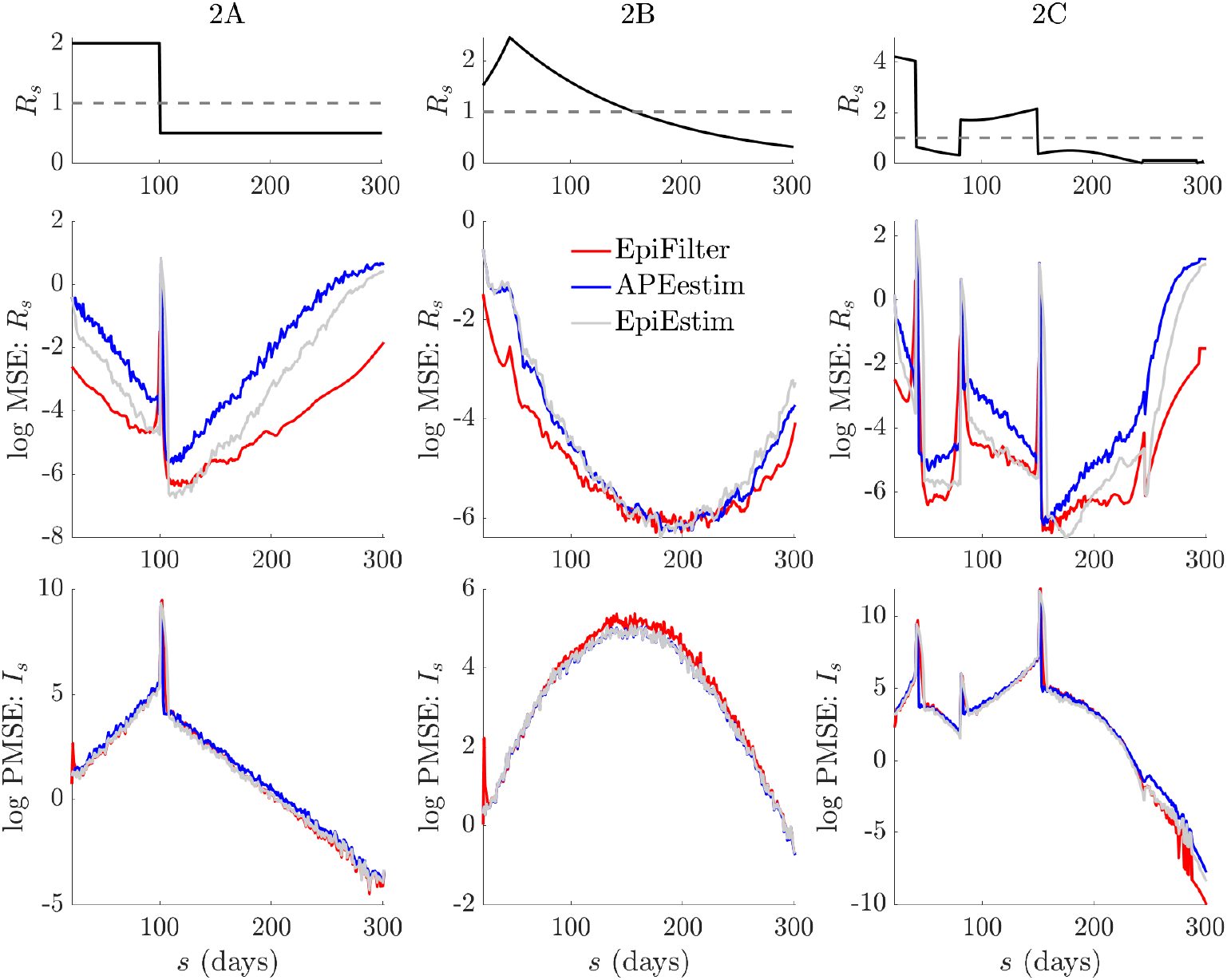
Temporal statistics of small or waning epidemics. We expand on the results from Fig. 2D by decomposing the MSE and one-step-ahead PMSE statistics across the 200 simulated trajectories for every scenario in Fig. 2. We do not consider the *k* = 31 EpiEstim example given its poor performance. We observe that EpiFilter significantly improves on MSE throughout the epidemic trajectory (and not only in periods of low incidence) while maintaining comparable prediction accuracies. Coverage statistics for these scenarios, which are given in the S1 Appendix (Fig. A3), confirm that EpiFilter also consistently contains the true *R*_*s*_ and *I*_*s*_ values within its credible intervals.

There the MSE and PMSE computed over the 200 replicate simulations are plotted with time. We observe a clear and significant reduction in MSE at most time points (both at low and large incidence) for EpiFilter, with similar PMSE performance, as compared to EpiEstim and APEestim. This confirms the benefits of smoothing solutions. Moreover, the coverage of both the true *R*_*s*_ and *I*_*s*_ of EpiFilter (i.e., the probability that *R*_*s*_ or *I*_*s*_ is contained within estimated or predicted 95% equal tailed credible intervals) is more consistent than all other approaches. This is shown in the S1 Appendix (Fig. A3). Thus, EpiFilter combines the advantages of APEestim and long-window EpiEstim and, further, is able to reliably detect transmission change-points automatically.

### Improved estimation between epidemic waves

Maintaining robust instantaneous reproduction number, *R*_*s*_, estimation when incidence, *I*_*s*_, becomes small is not just statistically important. Two possible outcomes may follow periods of small *I*_*s*_: either the epidemic goes extinct (elimination occurs, as in the previous section), or an additional wave of infection surfaces (resurgence) e.g., due to imports or unmonitored local transmission. Predicting which outcome is likely, in real-time, is of global concern as countries aim to relax interventions during the ongoing COVID-19 pandemic, while also minimising the risks of further resurgence [17], [39]. As changes in instantaneous reproduction numbers signal variations in transmission and hence incidence, reliably identifying and inferring *R*_*s*_ trends in the trough preceding potential new peaks can be crucial for preparedness, providing evidence for timely and effective epidemic interventions [10].

Reliable estimation of *R*_*s*_ between epidemic waves depends on the prior assumptions of the inference method used and on how that method relies on those assumptions when data are scarce [40], [41]. Here we examine this dependence and investigate cases where resurgence follows a low-incidence period. As in the above section, we compare EpiFilter (*η* = 0.1) with APEestim (optimal window *k*^∗^) and EpiEstim (weekly, *k* = 7, and monthly, *k* = 31, windows) over 200 simulated epidemics under the serial interval of Ebola virus. We explore scenarios depicting (A) epidemics that are initially controlled (*R*_*s*_ falls from 2.5 to 0.5 at *s* = 70) but which resurge just as quickly (*R*_*s*_ returns to 2.5 from *s* = 230), (B) periodic or seasonal transmission (*R*_*s*_ is sinusoidal with magnitude 1.3 ±1.2 and period of 120 time units) and (C) outbreaks with exponentially rising and then falling transmissibility (change-points at *s* = 40 and 190 and exponent rates 0.03, -0.015 and 0.02).

These examples are similar to some in [13] and describe diverse epidemics with multiple peaks and troughs. We provide representative runs of each scenario in A-C and collect MSE (for *R*_*s*_) and PMSE (relative to *I*_*s*_) distributions over the 200 runs of every scenario in D of Fig. 4. We also provide these statistics across time in Fig. 5. We observe a similar pattern in performance among the methods as in our previous analyses of Fig. 2 and Fig. 3. APEestim is best able to predict upcoming incidence and achieves the best PMSE as expected. While the monthly window EpiEstim is less useful in these cases (since prior reversion does not occur as often, though could be an issue for more extended troughs), the weekly window version loses predictive fidelity for minor MSE improvements.

**Fig. 4:**
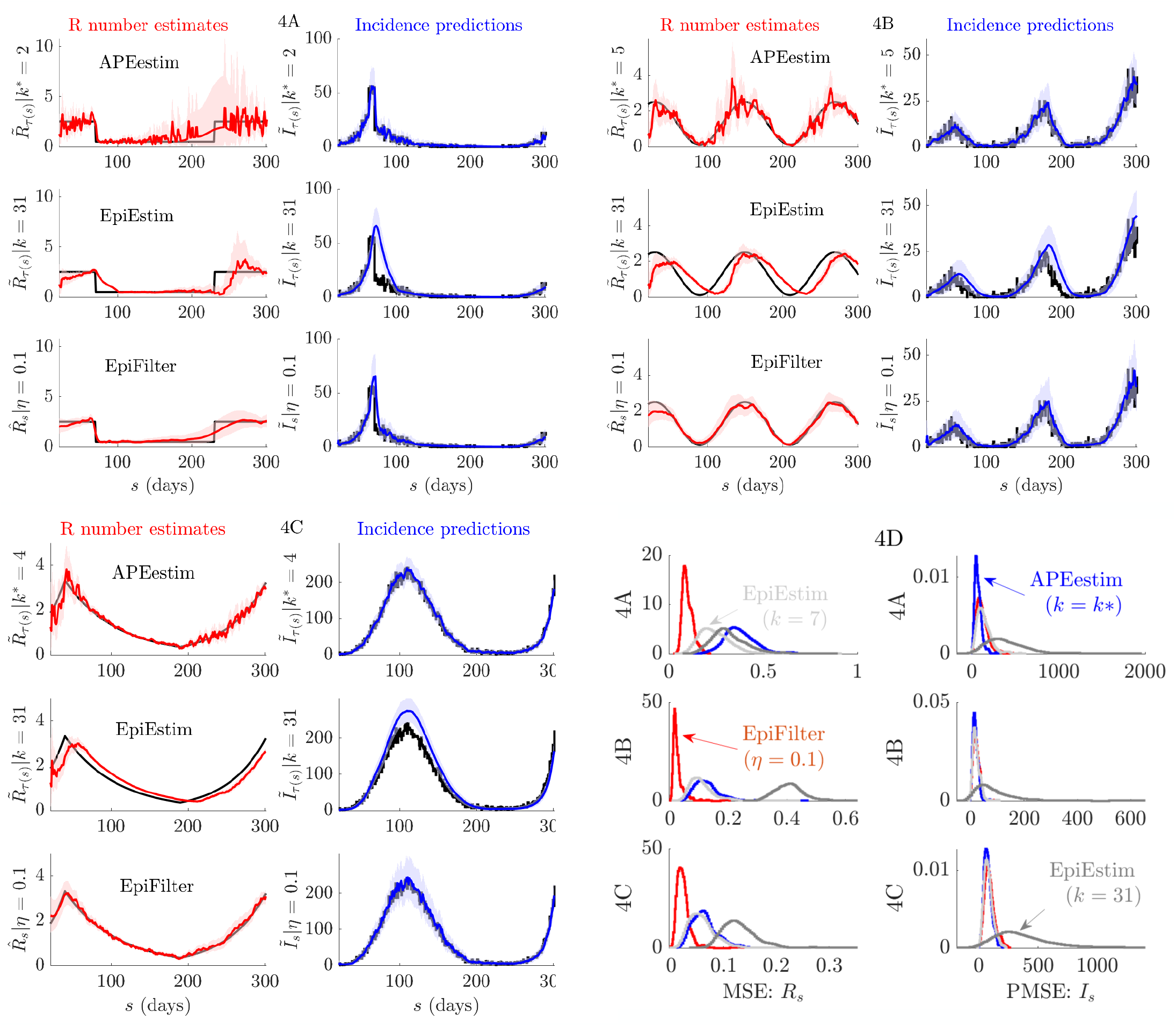
Epidemics with multiple waves. We compare reproduction number estimates (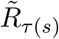 or 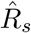) and one-step-ahead incidence predictions (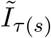 or 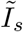) from APEestim with optimal window *k*^∗^, EpiEstim with window *k* and EpiFilter with state noise *η*. We simulate 200 epidemics with multiple waves of infection using the standard renewal model (Eq. (1)) for three scenarios, representative examples of which are given in A-C. The true *R*_*s*_ and *I*_*s*_ are in black. All mean estimates or predictions are in red and blue with 95% equal tailed credible intervals. APEestim and EpiEstim use a Gam(1, 2) prior distribution and EpiFilter a grid with *m* = 2000, *R*_min_ = 0.01 and *R*_max_ = 10. In D we provide statistics of the MSE of these estimates (relative to *R*_*s*_) and the PMSE of these predictions (relative to *I*_*s*_) for all 200 runs. We find EpiFilter is best able to negotiate troughs between epidemic peaks and hence infer resurging infectious dynamics, achieving significantly smaller MSE (2-10 fold reductions) and comparable PMSE to APEestim (which is optimised for prediction).

**Fig. 5:**
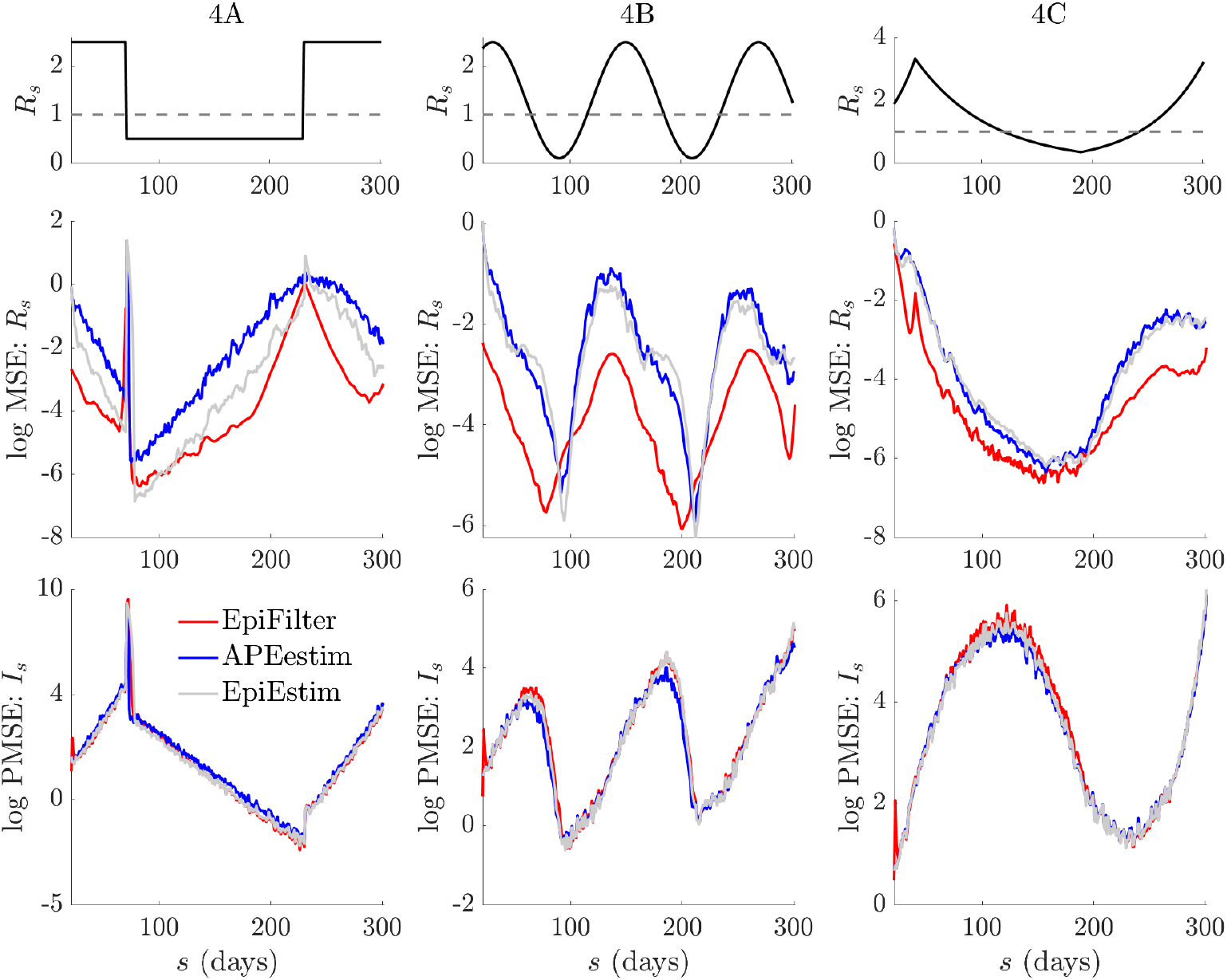
Temporal statistics of epidemics with multiple waves. We expand on the results from Fig. 4D by decomposing the MSE and one-step-ahead PMSE statistics across the 200 simulated trajectories for every scenario in Fig. 4. We do not consider the *k* = 31 EpiEstim example given its poor performance. We observe that EpiFilter significantly improves on MSE throughout the resurgent epidemic trajectory while maintaining comparable prediction accuracies. Coverage statistics for these scenarios, which are given in the S1 Appendix (Fig. A4), confirm that EpiFilter consistently contains the true *R*_*s*_ and *I*_*s*_ values within its credible intervals.

EpiFilter once again combines the advantages of the other approaches. For every scenario in Fig. 4 it provides accurate tracking of changes in *R*_*s*_ with stable credible intervals and a MSE that is at least 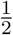 and sometimes even 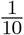 that of the next best method. The improvement in MSE is clear and maintained at almost every time point of each scenario, regardless of whether incidence is small or large. Concurrently, the incidence PMSE of EpiFilter rivals that of APEestim and it attains the most consistent coverage of the true *R*_*s*_ and *I*_*s*_ values, as shown in the S1 Appendix (Fig. A4). EpiFilter is therefore a powerful tool for detecting resurgence. We also find that the *η* = 0.1 parameter value seems to be an all-purpose heuristic, meaning that usage of EpiFilter can be simpler than EpiEstim and other window or change-point based methods. The improvements of EpiFilter likely result from its minimal assumptions (see Eq. (4)) and its increased information extraction. We next test our method on empirical data.

### COVID-19 in New Zealand and H1N1 influenza in the USA

The previous sections confirmed EpiFilter as a powerful inference and prediction tool, especially in data-poor conditions, using simulated epidemics. We now confront our method with empirical data from the 1918 H1N1 influenza pandemic in Baltimore (USA) [42] and the ongoing COVID-19 pandemic in New Zealand (up to 17 August 2020) [43]. The H1N1 dataset has been well-studied and so we first use this to benchmark EpiFilter. We clean this dataset by applying a 5-day moving average filter as recommended in [42]. Previous work [11] analysed this dataset with EpiEstim and found that sensible instantaneous reproduction number, *R*_*s*_, estimates are obtained when a weekly window (*k* = 7) is applied. However, more recent work, using APEestim [13], showed that while *k* = 7 provides stable estimates for this epidemic, it is a poor predictor of the incidence data. Instead, an optimised window of 2 days (*k*^∗^= 2) yields good predictions but the resulting *R*_*s*_ estimates are noisy.

We reproduce the instantaneous reproduction number estimates 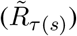 and incidence predictions 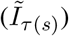 from both studies in Fig. 6 and compare them against EpiFilter with *η* = 0.1 (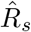 and 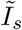). Top and middle rows of Fig. 6 illustrate the aforementioned trade-off between estimate stability and prediction accuracy. The bottom row confirms the power of EpiFilter. Our *R*_*s*_ estimates are of comparable stability to those of EpiEstim at *k* = 7, yet our prediction fidelity matches that of APEestim. Our improved inference again benefits from using more information (i.e., the backward pass in Fig. 1) and making less restrictive prior assumptions. We see the latter from the *R*_*s*_ credible intervals over 40 ≤ *s* ≤ 60. There EpiEstim seems overconfident, and this results in a rigid overestimation of incidence. However, EpiFilter mediates its estimate uncertainty to a level similar to APEestim.

**Fig. 6:**
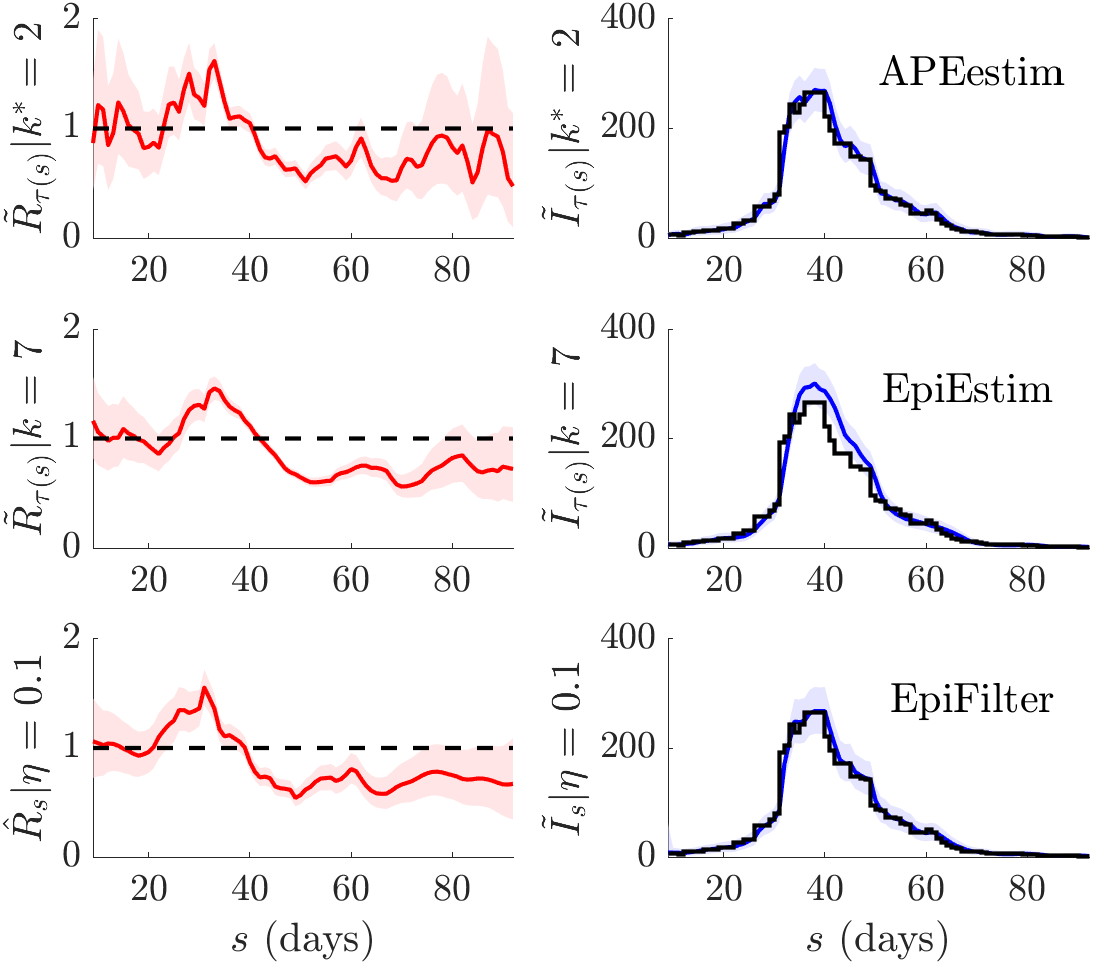
H1N1 influenza transmission in Baltimore (1918). We compare APEestim (top), EpiEstim with recommended weekly window (middle) (both with Gam(1, 2) prior distribution) and EpiFilter (with *m* = 2000, *η* = 0.1, *R*_min_ = 0.01 and *R*_max_ = 10) on the H1N1 influenza dataset from [42]. We use a 5-day moving average filter, as in [42], to remove known sampling biases. Estimates of reproduction numbers, *R*_*s*_, and corresponding 95% equal tailed credible intervals are in red. One-step-ahead predictions of incidence, *I*_*s*_, (with 95% credible intervals) are in blue with the actual incidence in black. We find that EpiFilter combines the benefits of APEestim and EpiEstim, achieving both good estimates and predictions.

We explore COVID-19 transmission patterns in New Zealand using incidence data up to 17 August 2020 from [43]. New Zealand presents an insightful case study because officials combined swift lockdowns with intensive testing to achieve and sustain very low incidence levels that eventually led to local elimination of COVID-19 [44]. However, an upsurge in cases in early August inspired concerns about a second wave (which led to new interventions and is why we do not consider data beyond 17 August). Here we investigate the time-varying transmission in New Zealand to see if this uptick suggests that the epidemic was resurfacing in mid-August. We believe smoothing can confer important inferential advantages in exactly these types of low incidence scenarios.

We make the common assumptions that case under-reporting is constant [11], which seems reasonable given the intensive surveillance employed by New Zealand [45]. We ignore reporting delays, which are known to be small [46] and use the COVID-19 serial interval distribution from [47]. We do not explicitly distinguish imported from local cases in our analysis. The latter could bias our study [28], [39] but our focus is on demonstrating differences between filtering and smoothing on instantaneous reproduction number, *R*_*s*_, trends and not on providing detailed *R*_*s*_ estimates during this period. We plot the results of our exploration in Fig. 7.

**Fig. 7:**
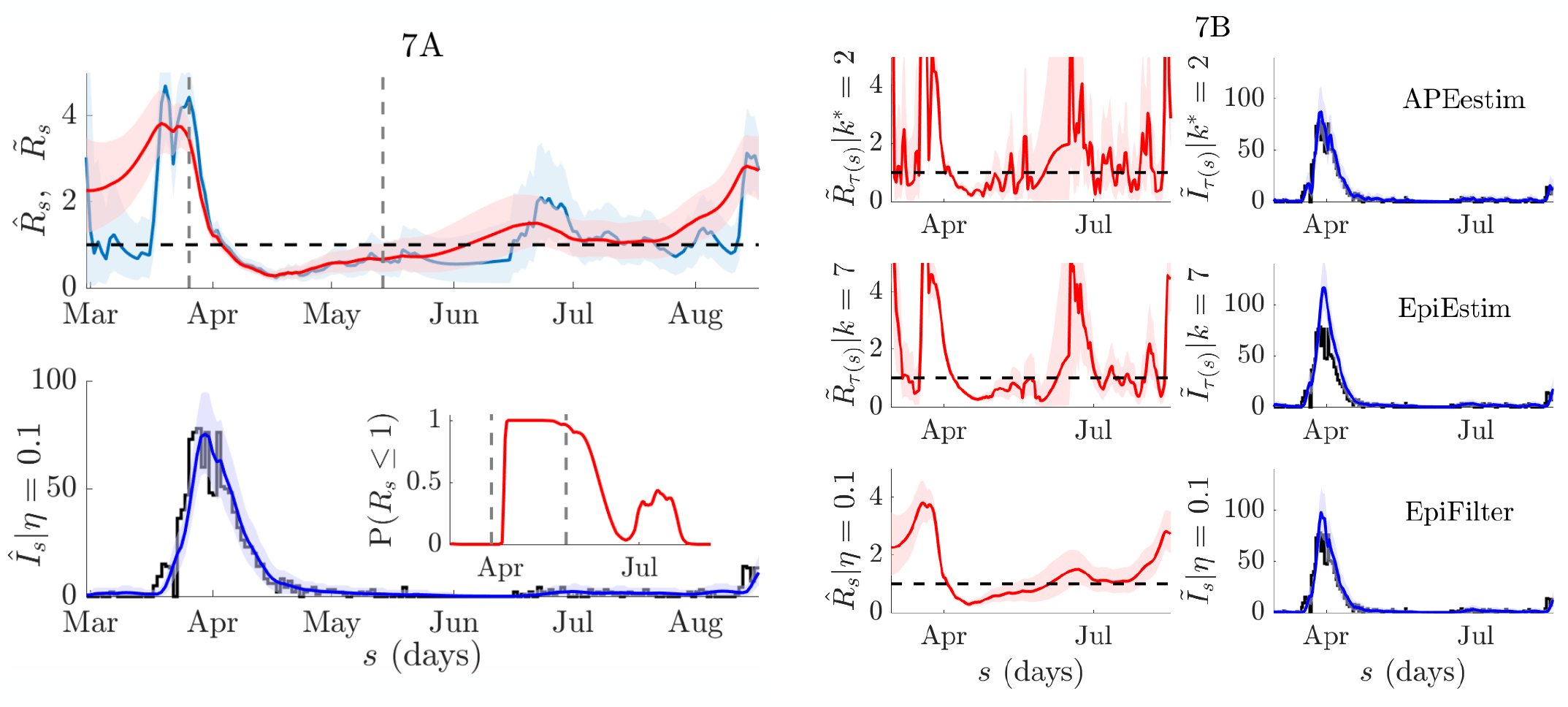
COVID-19 transmission in New Zealand. We compute smoothed and filtered reproduction number estimates, 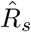 (red) and 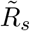 (blue) respectively, from the COVID-19 incidence curve for New Zealand (available at [43]) in the left panels. We use EpiFilter with *m* = 2000, *η* = 0.1, *R*_min_ = 0.01 and *R*_max_ = 10 with a uniform prior distribution over the grid ℛ. The top of 7A shows conditional mean estimates and 95% credible intervals for 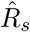 (red) and 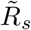 (blue). Vertical lines indicate the start and end of lockdown, a major intervention that was employed to halt transmission. The additional ‘future’ information used in smoothing has a notable effect. The bottom of 7A provides smoothed one-step-ahead predictions 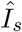 (blue, with 95% credible intervals) of the actual reported cases *I*_*s*_ (black). The inset gives the estimated probability of *R*_*s*_ ≤ 1. We observe a clear trend of subcritical transmission that eventually seeds a second wave by August. In 7B we compare EpiFilter with EpiEstim (using weekly windows) and APEestim (both with Gam(1, 2) priors) with all left subfigures presenting *R*_*s*_ estimates and right ones providing filtered *I*_*s*_ predictions. We observe that both APEestim and EpiEstim lead to largely unusable estimates that mask transmission trends, in sharp contrast to EpiFilter.

We apply EpiFilter and obtain filtered (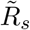, blue) and smoothed (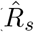, red) conditional mean estimates together with their 95% confidence intervals. These are in the left panels of Fig. 7 and computed from Eq. (5) and Eq. (6) respectively. The times of lockdown and release are included for reference. Interestingly, we see a notable difference in the quality of inference between 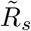 and 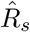. The former, as expected, is unreliable at the beginning of the incidence curve and features wider uncertainty and noisier trends. The smoothed 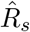, by using both forward and backward-looking data largely overcomes these issues and clarifies transmission dynamics. In the right panels we compare EpiFilter with EpiEstim (weekly windows) and APEestim. The difference is striking. Neither APEestim nor EpiEstim recovers a clear trend and both are appreciably worse than even the filtered estimate 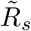.

Our smoothed analysis suggests that *R*_*s*_ has resurged and supports the re-implementation of measures around 14 August. We recover suppression of the initial wave in April, likely associated with the implementation of key interventions, including lockdown [46]. Following this, we infer a prolonged period of subcritical transmission, where ℙ(*R*_*s*_ ≤ 1) ≈ 1. Low and then zero incidence over this period strikingly destabilises both APEestim and EpiEstim and conceals the rise in *R*_*s*_ that occurs after July. EpiFilter signals this upsurge, which continually grows until August, where a second wave becomes likely. We also provide one-step-ahead predictions (which are from the smoothed 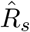 hence the notation 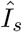) and their equal tailed 95% credible intervals against the reported incidence from [43]. These verify that 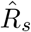 reasonably describes the data (we also present 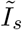 when comparing methods). We find that ℙ(*R*_*s*_ ≤ 1) ≈ 0 around July, further supporting this resurgence hypothesis.

While the analyses of the H1N1 and COVID-19 data above illustrate the advantages of EpiFilter, these estimates can be further improved. We used case data and not actual infection times (which relates to the approximation of the generation time by the serial interval distribution) and did not account for introductions, case ascertainment fractions and reporting delays. For more practical analyses it may be necessary to first compensate for these biases to obtain the best possible incidence curve. Methods from [6], [12], [48] can be applied if relevant incubation period, reporting distributions and contact tracing data are available to diagnose and correct for these issues. The resulting pre-processed incidence curve can then be input to EpiFilter to obtain more realistic *R*_*s*_ estimates.

## Discussion

Estimating time-varying trends in the instantaneous or effective reproduction number, *R*_*s*_, reliably and in real-time is an important and popular problem in infectious disease epidemiology [5]. As the COVID-19 pandemic has unfolded, the interest in solving this problem has only elevated with *R*_*s*_ playing a central role both in aiding situational awareness [8] and informing policymaking [49]. Initially, interest was on understanding how changes in *R*_*s*_ may correlate with interventions such as lockdowns and social distancing [9], [10]. However, as countries have entered waning phases of the pandemic and vaccine deployment has begun, focus has shifted to charactering how existing interventions can be relaxed with minimum risk [50]. The literature on intervention exit strategies is, however, still in development, and several challenges remain to modelling transmission.

One key challenge lies in understanding and inferring transmissibility during periods when the incidence of new cases is small [17]. Such periods may occur under sustained control measures and necessarily contain limited data, which make inferences difficult. Moreover, it is in these lulls that information on transmission may be crucial, helping to determine if the removal of interventions will lead to resurgence or if elimination is realistic by maintaining controls [28], [39]. While reproduction numbers are not the only analytic for assessing these outcomes, they do provide an important real-time diagnostic since upticks in *R*_*s*_ generally precede elevations in case loads. Unfortunately, current approaches to estimating *R*_*s*_ become underpowered, unstable or prior-constrained in these data-limited conditions [11], [26], [50]. These problems are only magnified when finer-scale analyses (where cases are fewer by division) are of interest (e.g., regional versus national level estimation).

In this paper we re-examined existing methodology for inferring instantaneous reproduction numbers, *R*_*s*_, from an engineering perspective. We observed that two of the most useful and popular inference approaches, EpiEstim [11] and the WT method (this computes cohort reproduction numbers, which are functions of *R*_*s*_) [2], only capitalise on a portion of the data available, deeming either upcoming or past incidence to be informative (see Fig. 1) [12]. This informative portion is directly controlled by prior assumptions on the speed of possible *R*_*s*_ changes, which are often characterised by a window of size *k*. Other methods also apply similarly strong change-point or state assumptions on *R*_*s*_, explicitly linking its variations with specific dates or events, for example [9], [26]. When data are scarce these assumptions can unduly control or skew inference.

In control engineering a common problem, known as filtering, involves optimally (in a MSE sense) estimating hidden Markov states, in real-time, from noisy and uncertain observations [18]. A related problem termed smoothing provides accompanying and optimal retrospective inferences [14]. By reinterpreting *R*_*s*_ as a Markov state (Eq. (4)) observed through a noisy renewal process (Eq. (1)) and defining *R*_*s*_ on a predetermined grid ℛ, we were able to construct exact filtering (Eq. (5)) and smoothing (Eq. (6)) solutions. This led to EpiFilter, which is our central contribution. Generally, filtering and smoothing can be involved and require sophisticated sequential Monte Carlo techniques [32]. However, because we make only minimal assumptions about *R*_*s*_, modelling it as a simple diffusion, we were able to solve these problems exactly and without complex sampling algorithms [29].

Our solutions are computationally simple, often executing in a few minutes (see S1 Appendix, Fig. A1), and deterministic i.e., precisely reproducible given the same data and settings. Our method replaces strong change-point or window size assumptions with one free parameter, *η*, which allows us to model some heterogeneity in transmission and sets the correlation among successive *R*_*s*_ values. We find that *η* = 0.1 serves as a general heuristic, providing good estimates and automatically detecting change-points and salient *R*_*s*_ dynamics over diverse scenarios (see Fig. 2, Fig. 4 and S1 Appendix, Fig. A2). This heuristic is also statistically justified by its good one-step-ahead predictive performance and consistent coverage of true simulated values (see S1 Appendix, Fig. A3-4) [13].

Importantly, EpiFilter is able to look both forward and backward through the incidence data, and so maximise the information extracted at every time point [34]. This property means it combines advantages from both EpiEstim and the WT method (see Fig. 1) and largely ameliorates their edge-effect issues [12]. These benefits, which also hold at large incidence, make EpiFilter a useful and robust tool for both real-time and retrospective *R*_*s*_ inference. We confirmed the advantages of EpiFilter by comparing it to EpiEstim and APEestim (a prediction optimised analogue to EpiEstim) on many simulated examples with periods of low incidence and epidemic resurgences (Fig. 2 and Fig. 4). Interestingly, we found EpiFilter was able to achieve significant 2-10 fold reductions in the MSE of *R*_*s*_ estimates without compromising predictive power or coverage of the true *R*_*s*_ and *I*_*s*_ values.

EpiFilter was especially better at negotiating periods of low incidence, offering a graceful degradation to its prior distribution or assumptions without sacrificing predictive accuracy. When incidence is low, it can be beneficial to use longer windows with EpiEstim [28]. This keeps *R*_*s*_ estimates reasonably stable but often leads to poor predictions [13]. APEestim, which optimises window size for prediction fidelity, showed that in many of the simulated scenarios short windows are necessary for describing transmission patterns. Consequently, we have a trade-off between estimate robustness and prediction accuracy. We found that EpiFilter overcomes this trade-off, concurrently achieving good estimates and predictions. In doing so, it revealed subcritical transmission trends and unmasked important signals of resurgence from noisy data in those periods.

We verified the practical utility and performance of EpiFilter on empirical data from the H1N1 pandemic of 1918 (see Fig. 6) and COVID-19 in New Zealand (see Fig. 7). In the first, which is a standard dataset that has been used to test previous *R*_*s*_ methods, we found that EpiFilter integrated the benefits of EpiEstim and APEestim to achieve simultaneously good estimates and predictions. A key use-case for EpiFilter is in signalling resurgence during low incidence. The COVID-19 epidemic in New Zealand featured precisely those dynamics [46]. While EpiEstim and APEestim were destabilised and unable to extract clear transmission trends, EpiFilter inferred subcritical *R*_*s*_ values and forewarned of resurgence by signalling an uptick in *R*_*s*_ just before a second wave become apparent. Recent, more involved COVID-19 analyses [39], have confirmed EpiFilter as a useful outbreak analytics tool.

Balancing the assumptions inherent to a model against the data it is applied on, to produce reliable inference is a non-trivial problem that is still under active investigation in several fields [15], [40], [41]. EpiFilter, by maximising the information extracted from available incidence data and minimising its state space model assumptions, appears to strike this balance as an estimator of instantaneous or effective reproduction numbers. Consequently, it performs strongly on a wide range of problems, including those involving sparse data, where other methods might struggle. Given its demonstrated advantages, straightforward formulation and theoretical underpinning, we hope that EpiFilter will be useful as a diagnostic tool for reliably signalling second waves of infection over multiple scales and more generally for assessing dynamical patterns in transmission both in real time and retrospectively. EpiFilter is freely available at https://github.com/kpzoo/EpiFilter.

## Data Availability

Code and data are available in both MATLAB and R at https://github.com/kpzoo/EpiFilter

## Acknowledgments

Thanks to Christl A Donnelly for thoughtful and helpful comments.

