## supplementary information for "Improved estimation of time-varying reproduction numbers at low case incidence and between epidemic waves"

Kris V Parag†, [1]

[1]MRC Centre for Global Infectious Disease Analysis, Imperial College London, London, W2 1PG, UK

†

### VALIDATING THE MECHANICS AND PERFORMANCE OF EPIFILTER

Having constructed EpiFilter (see Methods) we now investigate its mechanics and performance. First we show how the choice of grid resolution $m$ affects the accuracy and computational complexity of our smoothed instantaneous reproduction number estimates, $\hat{R}_s$. Our recursive implementation is also known as a grid smoother, since its estimates are optimal (in a MSE sense) but conditioned on the grid $\mathcal{R}$ [1], [2]. In the left panels of Fig. A1 we reconsider the empirical H1N1 and COVID-19 examples, from the main text, at various $m$. We find that EpiFilter has linear complexity in $m$ but that $\hat{R}_s$ (averaged over 50 replicate runs) converges quickly, both in mean and variance (latter not shown). Execution times, which include both $R_s$ estimation and incidence ($I_s$) prediction, are only a few minutes, generally. While the $R_s$ smoother is always fast, the $I_s$ prediction block may be slower for very large epidemics (e.g., 1000s of cases per day), resulting in $\sim 10$ minute run times.

As a rule-of-thumb setting $m \geq 10^3$ safely guarantees convergence, and still executes in a few minutes. Note that estimates at small $m$ are still valid. They represent coarser assumptions on reproduction numbers and provide minimised MSE estimates under those assumptions. We illustrate a coarse estimate and how it converges for a simulated example in the right panels of Fig. A1. There, we also test the behaviour under different choices of the process or state noise parameter $\eta$. This parameter is vaguely similar to the window size parameter, $k$, in EpiEstim approaches, but is easier to select. It controls the autocorrelation among successive $R_s$ and allows some modelling of heterogeneity (hence the term state noise). Fig. A1, shows how too small $\eta$ might lead to underfitting, while overly large $\eta$ (where we would not consider $\eta \geq 1$ given Eq. (4b) of the main text) could promote overfitting.

However, while the optimal window size in EpiEstim varies with the form of the true $R_s$ (e.g., large $k$ is needed for stable $R_s$ time-series and small $k$ for strongly fluctuating ones [3]), we find that a fixed $\eta = 0.1$ works well across diverse epidemic scenarios and enables automatic detection of transmission change-points. This is an advantage over approaches requiring explicit change-point modelling [4]. We examine this further in Fig. A2. There we compare $R_s$ estimates from APEestim (with optimal window $k$) [3] to EpiFilter ($\eta = 0.1$, $m = 2000$) for simulated epidemic scenarios. We generate epidemics via the renewal model (Eq. (1) of the main text) using the Ebola virus serial interval distribution from [5] and ensure the prior distributions of both methods have the same support. The epidemics simulated span the range of possibilities (e.g., they may have large or small incidence).

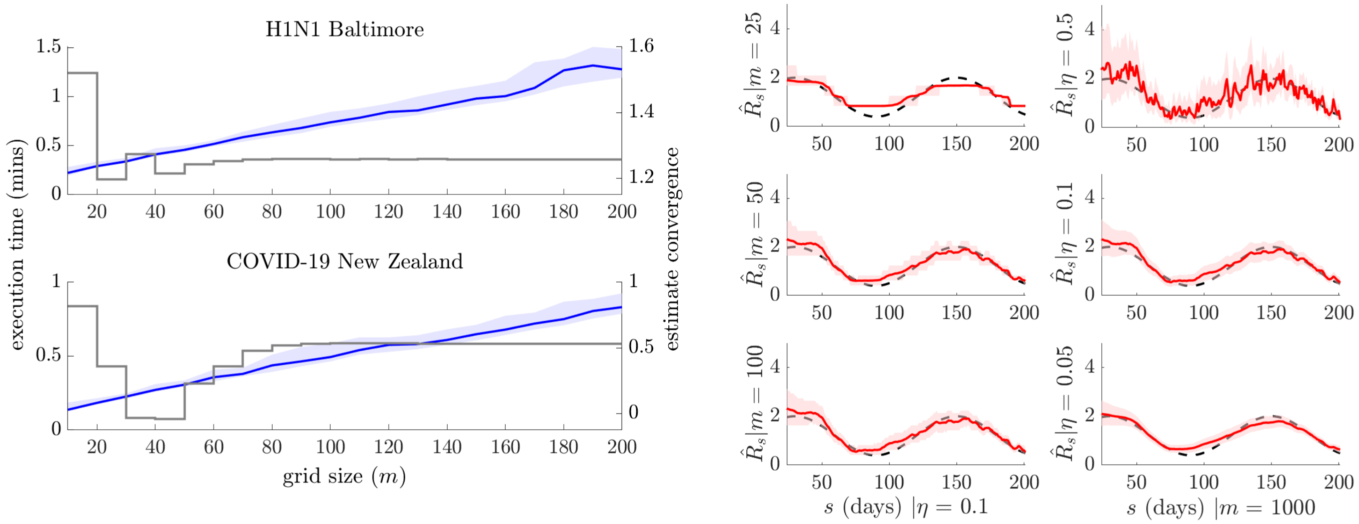


Fig. A1: **EpiFilter mechanics.** We examine the computational complexity and accuracy of EpiFilter. Left panels indicate how increasing grid resolution, $m$, leads to a linear increase in execution time (blue with 95% credible intervals) for the empirical case studies of the main text. These times are averaged over 50 replicate runs (using the R implementation) and include the latency in computing both reproduction number, $R_s$, estimates and incidence, $I_s$, predictions. The mean estimate of $R_s$, averaged over time and the replicates, is in grey, and converges quickly. The variance of these estimates, while not shown, converges equally fast. Right panels provide an illustrative example of how estimates $\hat{R}_s$ (red with 95% credible intervals) change with $m$ and choices of process noise parameter, $\eta$. Even at coarse grid sizes (small $m$) the estimates are sensible. Reducing $\eta$ sharpens confidence intervals but can potentially underfit. Increasing $\eta$ adds state noise. We find that $\eta = 0.1$ serves as a good general heuristic. All simulations are at $R_{\text{min}} = 0.01$ and $R_{\text{max}} = 10$ and the true $R_s$ (a seasonal epidemic) is in black.

We find EpiFilter outperforms APEestim, despite our using a single $\eta$ value, with improvements especially stark for the simulations with subcritical transmission (as detailed in main text). This single $\eta$ contrasts the discordant optimal $k$ values and allows EpiFilter to recover the salient dynamics for epidemics with stable trajectories, seasonal variations and rapidly or gradually fluctuating transmission. In none of these scenarios does the true $R_s$ have either the state model of Eq. (4b) of the main text or the sliding-window relationship in EpiEstim. Hence, these simulations also indicate robustness of the different inherent state assumptions in both approaches to moderate model mismatch. We use single runs to showcase how a practical epidemic would be processed in real-time. Neither method depends on random algorithms (e.g., MCMC) so their estimates will remain constant under the same parameters.

Last, we complete our batch analysis of EpiFilter performance by examining coverage statistics. In Fig. 3 and Fig. 5 of the main text a range of waning and resurging epidemic examples were explored. There we found that EpiFilter considerably improves the MSE when inferring instantaneous reproduction numbers, $R_s$, relative to EpiEstim and APEestim. APEestim, expectedly, had the best PMSE when predicting incidence, $I_s$, but EpiFilter was only marginally worse. In Fig. A3 and Fig. A4 we plot the coverage (over the 200 runs of each scenario from Fig. 3 and Fig. 5) against time i.e., the probability that the $R_s$ estimate or $I_s$ prediction equal tailed credible intervals of a given method contain the true $R_s$ and $I_s$ values from the simulations. We find that EpiFilter generally

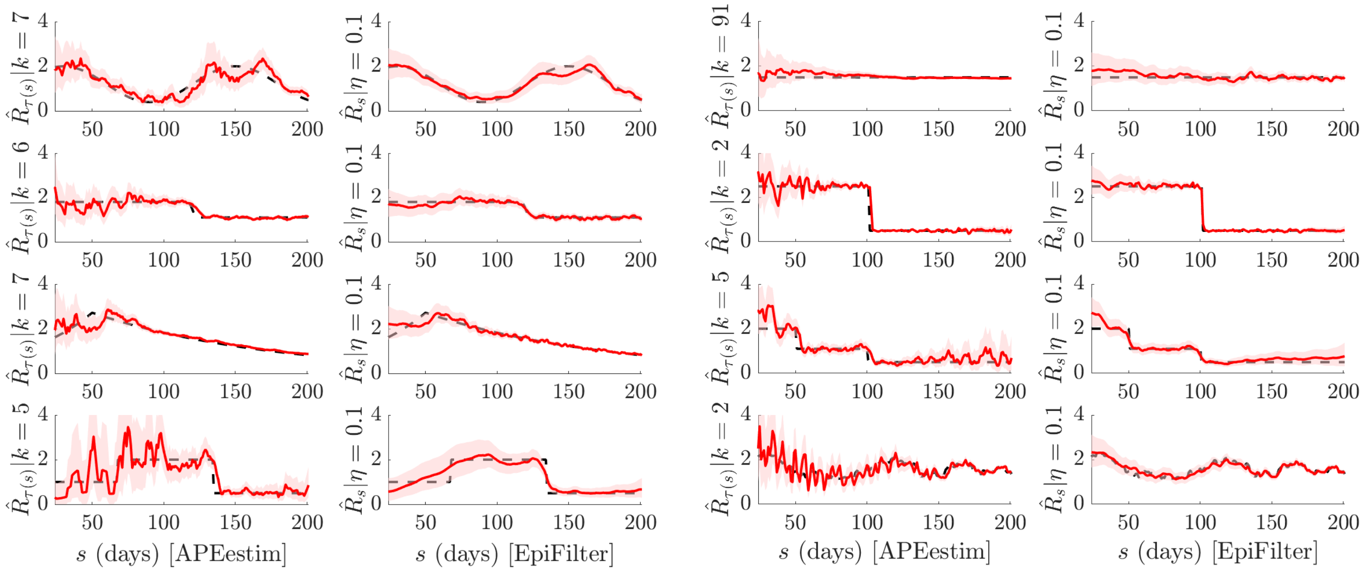


Fig. A2: **Validation against APEestim.** We compare estimates from EpiFilter (right panels) with optimised ones from APEestim (left panels [3]) over various general epidemic scenarios (which may have large or small incidence). These are simulated using Eq. (1) of the main text and not restricted to epidemics that may face elimination or resurgence as in Fig. 2 and Fig. 4 of the main text. We use $m = 2000$, $R_{\text{min}} = 0.01$ and $R_{\text{max}} = 10$ for EpiFilter. To be fair we use a $\text{Gam}(1, 2)$ prior distribution with APEestim, which sums to 1 by $R_{\text{max}}$. Here $k$ is the optimal window choice in APEestim (under predictive error) and $\eta = 0.1$ is fixed for EpiFilter inferences. True $R_s$ is in black and conditional mean estimates with 95% confidence intervals are in red. EpiFilter is more stable, smoother and minimises estimate uncertainty in all scenarios. While the optimal $k$ changes considerably among epidemics, $\eta = 0.1$ is able to automatically detect salient changes in transmission for all examples considered.

has the most consistent coverage across all scenarios for both $R_s$ and $I_s$, confirming its performance.

### Acknowledgments

Thanks to Christl A Donnelly for thoughtful and helpful comments.

### Funding

This work is jointly funded under grant reference MR/R015600/1 by the UK Medical Research Council (MRC) and the UK Department for International Development (DFID) under the MRC/DFID Concordat agreement and is also part of the EDCTP2 programme supported by the European Union.

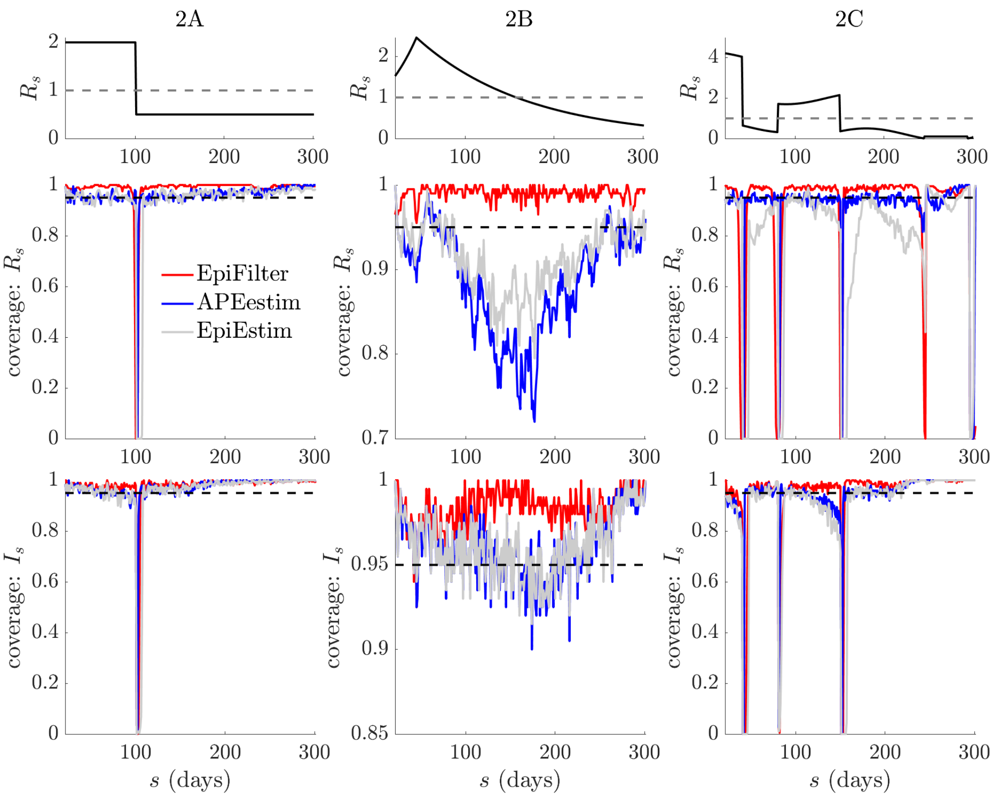


Fig. A3: **Coverage statistics of waning epidemics** We provide the probability that the true values of instantaneous reproduction numbers, $R_s$, and incidence, $I_s$, from each scenario in Fig. 3 of the main text (over 200 runs) are contained within the equal tailed 95% estimate and prediction credible intervals generated by EpiFilter ($\eta = 0.1$), APEestim ($k = k^*$) and EpiEstim (weekly, $k = 7$ windows). EpiFilter provides the most consistent coverage.


[4] S. Flaxman, S. Mishra, A. Gandy, *et al.*, "Estimating the effects of non-pharmaceutical interventions on COVID-19 in Europe," *Nature*, vol. 584, pp. 257–261, 2020.

[5] M. Van Kerkhove, A. Bento, H. Mills, *et al.*, "A review of epidemiological parameters from Ebola outbreaks to inform early public health decision-making," *Sci. Data*, vol. 2, p. 150019, 2015.

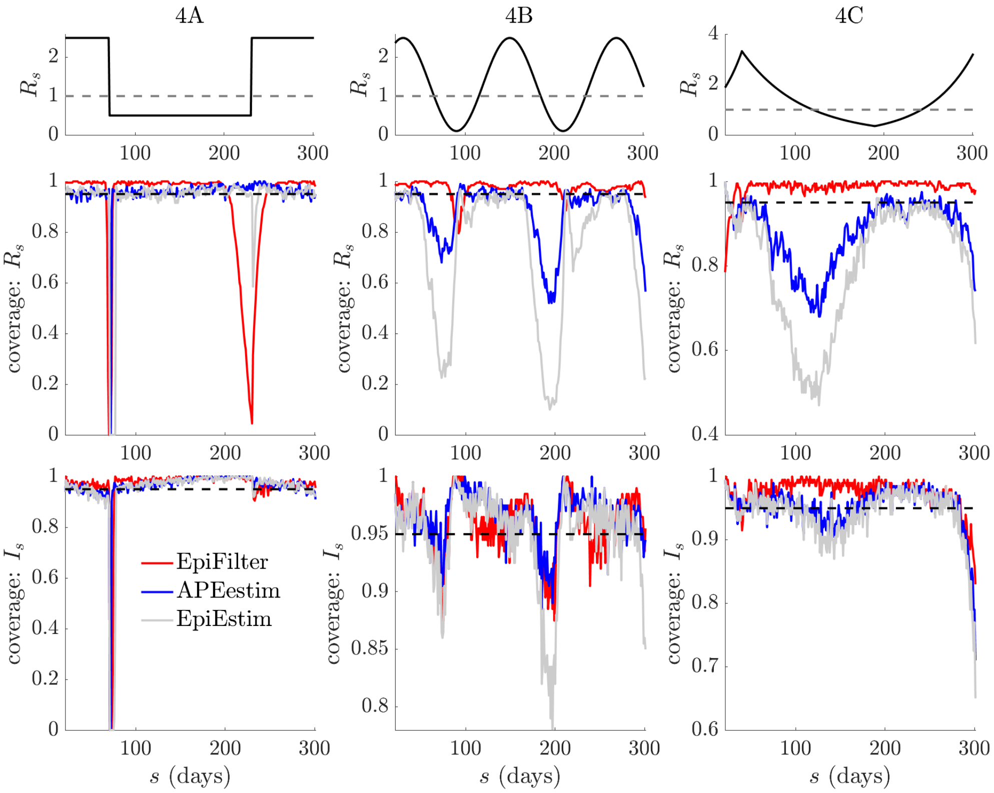


Fig. A4: **Coverage statistics of resurging epidemics.** We provide the probability that the true values of instantaneous reproduction numbers, $R_s$, and incidence, $I_s$, from each scenario in Fig. 5 of the main text (over 200 runs) are contained within the equal tailed 95% estimate and prediction credible intervals generated by EpiFilter ($\eta = 0.1$), APEestim ($k = k^*$) and EpiEstim (weekly, $k = 7$ windows). EpiFilter provides the most consistent coverage.
